# Domain specific models outperform large vision language models on cytomorphology tasks

**DOI:** 10.1101/2025.05.05.25326989

**Authors:** Ivan Kukuljan, Muhammed Furkan Dasdelen, Julia Schäfer, Michele Buck, Katharina S. Götze, Carsten Marr

**Affiliations:** Institute of AI for Health, Computational Health Center, Helmholtz Munich, Ingolstädter Landstr. 1, 85764 Neuherberg, Germany; Technical University Munich, School of Medicine and Health, Ismaninger Str. 22, 81675 München, Germany; Klinikum rechts der Isar, III. Medizinische Klinik, Technische Universität München, Ismaninger Str. 22, 81675 München, Germany

**Author notes:** Shared first authorship (Ivan Kukuljan, Muhammed Furkan Dasdelen).

## Abstract

Large vision-language models (LVLMs) show impressive capabilities in image understanding across domains. However, their suitability for high-risk medical diagnostics remains unclear. We systematically evaluate four state-of-the-art LVLMs and three domain-specific models on key cytomorphological benchmarks: peripheral blood cell classification, morphology assessment, bone marrow cell classification, and cervical smear malignancy detection. Performance is assessed under zero-shot, few-shot, and fine-tuned conditions. LVLMs underperform significantly: the best LVLM achieves a zero-shot F1 score of 0.057 ± 0.008 for malignancy detection—near random (0.039)—and only 0.15 ± 0.01 in few-shot. In contrast, domain-specific models reach up to 0.83 in accuracy. Even after fine-tuning, a dedicated hematology model outperforms GPT-4o. While LVLMs offer explainability via text, we find the visual-language grounding unreliable, and the morphological features mention by the model often do not match the single cell properties. Our findings suggest that LVLMs require substantial improvements before use in high-stakes diagnostic settings.

**Key findings:**

- LVLMs perform poorly on cytomorphology tasks, often near chance level and far below domain-specific models.
- Even after fine-tuning, LVLMs lag behind domain-specific models.
- While LVLMs provide textual justifications, these often reflect generic descriptions rather than image-specific morphological features.

## Introduction

With the ability to process multimodal data, large vision-language models (LVLMs) have emerged as promising tools for medical image analysis, disease classification, and report generation^1–5^. Several studies have assessed the performance of LVLMs on medical imaging tasks: Jiang et al. evaluated LVLMs on endoscopy, chest X-ray, and skin lesion images^6^, while Royer et al. conducted a broader evaluation of medical LVLMs across multiple datasets, including ultrasound, mammography, radiology, dermatology, microscopy, and other general medical tasks^7^. Hartsoch and Rasool^8^ and Jeong et al.^9^ assessed LVLMs for medical report generation and question answering, whereas Wu et al. focused on their performance in radiology tasks^10^. Additionally, Tu et al. evaluated Med-PaLM-Multimodal on a wide range of medical imaging datasets, spanning pathology, radiology, and mammography^11^.

Despite this growing interest in medical LVLMs, their performance in cytomorphology remains largely unexplored. Cytomorphology is a cornerstone of hematology, pathology, and gynecology, offering cost-effective, minimally invasive diagnostics^12,13^. At the same time, it presents an ideal use case for AI model development and evaluation, benefiting from extensive annotated datasets and 180 years of morphological knowledge accumulated since Virchow’s first systematic cellular diagnoses^14^. Yet, the field faces a global shortage of trained cytologists—even in high-income countries—highlighting a potential role for artificial intelligence (AI) to support diagnostic workflows^15^. Prior work has shown that specialized models can achieve high performance on cervical smear screening^16–19^, blood cell classification^20–23^, and leukemia diagnosis^24–28^, with recent efforts developing foundation models for cytomorphology^29^.

Here, we evaluate the performance of LVLMs to answer the following questions: How well do generalist and medical-specific LVLMs perform on cytomorphology tasks? Is it more efficient to fine-tune LVLMs for cytomorphology-specific tasks, or is it better to develop dedicated AI models? Can we trust textual explanations these models provide? We systematically evaluated generalist LVLMs (GPT-4o, Gemini 2.0 Flash, LLama-3.2, DeepSeek-VL2), as well as medical-specific models (LLaVA-Med, CONCH, BiomedCLIP) on a diverse set of cytomorphology datasets, assessing their zero-shot and few-shot performance. Moreover, we fine-tuned GPT-4o for peripheral blood cell classification and compared its performance with a hematology foundation model.

## Methods

### Datasets & tasks

To evaluate the performance of the most important LVLMs on cytomorphology tasks, we selected four data sets for classifying cell types or lesion types (peripheral blood cells, bone marrow cells and cervical cells) and one dataset quantifying peripheral blood cell morphologies (Fig. 1A):

- **HiCervix**^30^ comprises 40,229 cervical cells from 4,496 whole slide images, categorized into 29 classes. The HiCervix dataset includes normal epithelial cells, infectious agents, and malignant cells.
- **Acevedo** et al.^31^ provide 17,092 white blood cell images from peripheral blood smears, labeled with 11 different cell type annotations.
- **BMC**^20^ is a collection of 171,373 white blood cell images from bone marrow smears collected from 945 patients. The cells were expert-labeled into 21 different cell types.
- **WBCAtt**^32^ contains morphology annotations for 10,300 images from the Acevedo data set^31^. Labels are provided for 11 fine-grained morphological attributes like nucleus shape, chromatin density, granularity, or cytoplasm texture.
- **MLL23**^33^ was used only as an external test set for fine-tuned models. It includes over 40,000 expert annotated peripheral blood single cell images categorized into 18 classes.

**Fig. 1.**
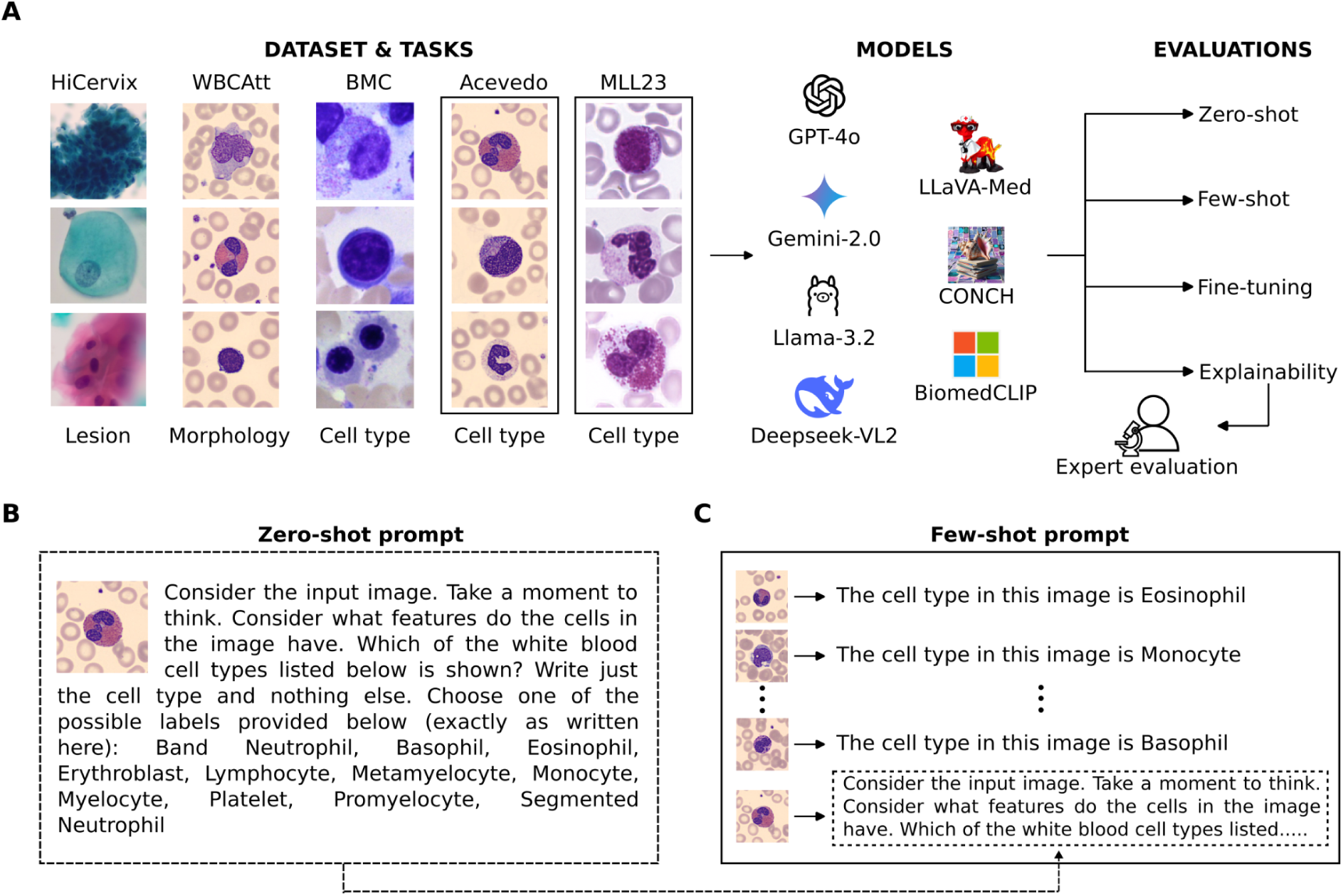
Systematic evaluation of Large Vision Language Models (LVLMs) in cytomorphology. **A.** We use five cytomorphology and hematology datasets with respective tasks (HiCervix: cervical smear lesion classification; WBCAtt: white blood cell morphology classification; BMC: bone marrow cell type classification; Acevedo: peripheral blood cell type classification, MLL23: out-of-domain peripheral blood cell type classification) to assess generalist LVLMs (GPT-4o, Gemini-2.0, Deepseek-VL2, LLaMA-3.2) alongside domain-specific models (LLaVA-Med, CONCH, BiomedCLIP) under zero-shot, few-shot, and fine-tuned conditions. **B.** In zero-shot evaluation, the prompt consists of a single image and a corresponding text query. **C.** In few-shot evaluation, the prompt includes an example image from each class with the corresponding label, followed by the input image and the text query.

For each dataset, we randomly sampled 50 images per class (or the maximum available samples if fewer than 50 images were present) as a test set across zero-shot, few-shot, and fine-tuning experiments. For few-shot learning, we selected an independent training set containing one image per class (Fig. 1C). For fine-tuning, only the Acevedo data set was used. We selected an independent subset of 200 images per class for training, and a separate validation set of 50 images per class. MLL23 was used exclusively for out-of-domain evaluations to assess the generalizability of fine-tuned models. The MLL23 test set contained only the classes present in Acevedo to ensure a fair comparison.

### Models

We evaluated four leading large vision-language models:

- **GPT-4o**^34^ is the Flagship model by OpenAI that can reason across audio, vision, and text in real time. The authors do not disclose the model’s architecture nor size.
- **Gemini-2.0-flash-exp**^35^ is Google Deepmind’s flagship vision language model. The authors do not disclose the model’s architecture nor size.
- **Llama-3.2-multimodal-11B**^36^ is the smaller of Meta AI’s vision language models with 11 billion weights, runnable on a single NVIDIA A100 GPU.
- **Deepseek-VL2-small**^37^ belongs to the latest 2nd generation of DeepSeek vision language models. We evaluate the small versions with 2.8 billion weights.

We also evaluated three models specifically designed for the biomedical domain:

- **LLaVA-Med**^38^ is Microsoft’s vision language model for biomedical images. It has been trained on 15 million biomedical image-text pairs.
- **CONCH**^39^ is a state-of-the-art vision language foundation model for computational pathology, trained on over 1.17 million image-caption pairs.
- **BiomedCLIP**^40^ is a biomedical vision-language foundation model pretrained on the 15 million image-text pairs.

Additionally, we included a hematology-specific model in our fine-tuning experiments for comparison with GPT-4o:

- **DinoBloom**^29^ is the state-of-the-art hematology foundation model. It is based on DINOv2 and trained on over 380,000 white blood cell images. We use the DinoBloom-S version with 22 million weights.

GPT-4o and Gemini-2.0 models are only available commercially through API calls, while the other models can be downloaded and run locally.

### Evaluations

#### Zero-shot

The model was presented with an image and asked to classify the cell it contained (Fig. 1B). Its prediction was then compared to the ground truth label from the dataset. The model was provided with a predefined list of labels from which it could choose. To increase the performance of the model, we instructed the model to take a moment to analyze the cell’s features before making a decision. To ensure a definitive classification, we removed ambiguous categories such as “not clear” or “not identifiable” from the list of possible answers. We evaluated zero-shot performance across all datasets. The prompt for the Acevedo dataset is shown in Fig. 1B. For the zero-shot CONCH and BiomedCLIP model evaluations, we compared embedding similarities between the image and the prompt text (see Supplementary Methods). The variance of the scores was computed by splitting the models’ answers into five non overlapping folds and computing the scores for each.

#### Few-shot

The model was first shown one example image for each cell class in the dataset, along with a description of the corresponding class (Fig. 1C). It was then presented with an unknown image and asked to classify it. As in zero-shot evaluation, the model selected from a predefined list of possible answers. We evaluated few-shot performance across all datasets.

#### Fine-tuning

To assess how effectively LVLMs could learn to interpret cytomorphology images, we fine-tuned GPT-4o via API access using the training subset of the Acevedo dataset. The same test subset of Acevedo dataset was used as in the other evaluations (non overlapping with the train set). The number of fine-tuning samples per class was n = 1, 5, 10, 25, 50, 100, and 200. We compared the fine-tuned GPT-4o with the fine-tuned multilayer perceptron (MLP) on top of the DinoBloom model at each fine-tuning sample size. We also included MLP on top of the DINOv2—a non-medical-domain pretrained model—as a baseline. Separate models were trained for each dataset size. We specifically chose the Acevedo blood cell dataset for fine-tuning, as DinoBloom had not been trained on this dataset, ensuring a fair comparison between the two models. We also tested the fine-tuned models on the external test set, MLL23.

#### Answer cleaning

Although the models were instructed to provide a short answer containing the cell class only, they occasionally generated longer responses with explanations. To address this, we processed the answers through a chatbot once more, asking it to extract only the cell class from the chatbot’s answer. For consistency and reliability, GPT-4o was used for this, ensuring uniform conditions across all models.

### Explainability

We evaluated explainability of the four generalist models (GPT-4o, GPT-4o fine-tuned with 200 images per class, Gemini-2.0, and Llama-3.2) on the Acevedo data set.

#### Quantitative feature importance

We presented the model with all 549 cells in the Acevedo test set, one by one, and asked it to classify the cell. Then it was presented with a list of 19 morphological features (see Fig. 4A) and asked to assign them a score on how relevant they were for the classification with the following prompt:

*“Consider the input image. […] Which of the white blood cell types listed below is shown? […] Now consider the cell features listed below. Think how much each of them contributed to your cell classification decision that you made above. Next to each feature, write an importance score how much the feature was important for your classification decision. The scores should be float numbers. All the scores together should sum to 100*.

*Cell Shape, Cell Size, Nuclear Shape, Nuclear Segmentation, Nuclear-to-Cytoplasmic Ratio, Nuclear Membrane Appearance, Nucleoli, Chromatin Pattern, Cytoplasmic Volume, Cytoplasmic Color, Cytoplasmic Border, Granule Presence, Granule Type, Inclusions (Presence of Auer rods, Döhle bodies, or other cytoplasmic inclusions), Cytoplasmic Basophilia, Erythrocytes, Platelets, Thrombocytes, Surrounding of the cell, Technical properties of the image (resolution, light, noise, etc.). Are there any other features that you consider important for the classification decision? If yes, write them below.”*

We averaged the scores for all the cells and for individual cell types (Fig. 4A) for those answers where the predicted label was true.

#### Model explanation

For the Acevedo test set, we asked the model to classify a cell and then provide a free-text explanation of the decision (Fig. 4BC) using the following prompt: *“Consider the input image. […] Which of the white blood cell types listed below is shown? […] Explain in detail your decision and the reasoning that lead you to the decision. Which parts of the cells and features did you consider? How certain are you about your classification? Which other labels could be correct? Why did you choose this label in the end?”*

#### Expert evaluation

We randomly selected 10 images per cell type (5 correctly and 5 incorrectly classified images), along with their corresponding explanations generated by the fine-tuned GPT-4o. All images were de-identified and presented to an expert cytomorphologist (M.B.) without indicating whether they were correctly or incorrectly classified. The expert was first asked to classify each image independently, and then provided with the corresponding GPT-4o-generated explanation. The expert rated each explanation based on how well it aligned with the cell’s morphological characteristics, using a 5-point scale: 1 – excellent, 2 – good, 3 – fair, 4 – poor, 5 – misleading or completely incorrect.

#### Computer vision

To assess the model’s understanding of cellular components, we asked it to highlight the nucleus in blue, granules in green, the entire white blood cell in pink, and red blood cells in purple, one at a time. We also asked the model to highlight parts of the cell relevant for the cell classification. API access could not be utilized, as it currently lacks image generation capabilities. Gemini 2.0 failed to generate meaningful images. Therefore, we focused our evaluation on GPT-4o.

## Results

We evaluated four generalist large vision-language models (LVLMs) and three medical domain-specific LVLMs on four different data sets (Fig. 1) and quantified their performance using the weighted F1 score (Table 1). As a reference, we considered the performance of models reported in the original papers of the respective dataset. Weighted F1 performance of a random model was calculated based on class distribution. LVLMs perform very poorly on zero-shot cell classification tasks, often yielding results similar to random performance and consistently far below the performance of models specifically designed for these tasks. For instance, the highest LVLM score for bone marrow cell classification was 0.094±0.018 (achieved by Gemini-2.0), compared to 0.049 for random guessing, and 0.75 for the model reported in the original study (Table 1). Similarly, for the cervical smear dataset, Gemini-2.0 achieved the best score of 0.057±0.011, while the random model scores 0.039, and the reference model reached an average accuracy of 0.83. Notably, GPT-4o frequently stated that it did not know the answer to this task, likely due to the scarcity of publicly available cervix cytomorphology datasets.

**Table 1.** Large visual language models exhibit near-random performance in zero-shot and few-shot evaluations on four cytomorphology tasks. The weighted F1 scores achieved by the LVLMs are far from the state-of-the-art results from the literature (last row), and close to random performance. Few-shot learning doubles the scores for Gemini-2.0 and GPT-4o in almost all tasks, however, they are still far below the state of the art. Mean and standard deviation were computed by splitting the test set into five balanced folds. Results from the original publication are α: averaged accuracy, β: accuracy, γ: weighted F1 score, and δ: averaged macro F1 score, as listed in the original publication.

|  | **HiCervix** | **Acevedo** | **BMC** | **WBCAtt** |
|---|---|---|---|---|
| **Random** | 0.038 | 0.091 | 0.047 | 0.36 |
| **Zero-shot performance (weighted F1)** |  |  |  |  |
| Gpt-4o | 0.035±0.015 | 0.22±0.02 | 0.087±0.018 | 0.35±0.03 |
| Gemini-2.0 | **0.057±0.008** | **0.25±0.04** | **0.094±0.015** | **0.49±0.02** |
| Llama-3.2 | 0.018±0.005 | 0.11±0.03 | 0.042±0.015 | 0.36±0.02 |
| Deepseek-vl2 | 0.0079±0.0025 | 0.015±0.001 | 0.0051±0.0008 | 0.26±0.02 |
| LLaVA-Med | 0.015±0.011 | 0.024±0.009 | 0.023±0.007 | 0.38±0.01 |
| CONCH | 0.0086±0.0023 | 0.031±0.007 | 0.030±0.001 | 0.25±0.01 |
| BiomedCLIP | 0.030±0.01 | 0.078±0.013 | 0.030±0.01 | 0.35±0.02 |
| **Few-shot performance (weighted F1)** |  |  |  |  |
| Gpt-4o | 0.088±0.015 | 0.36±0.02 | 0.12±0.02 | 0.64±0.03 |
| Gemini-2.0 | **0.15±0.01** | **0.56±0.04** | **0.20±0.02** | **0.68±0.02** |
| Llama-3.2 | 0.011±0.008 | 0.036±0.009 | 0.021±0.007 | 0.39±0.01 |
| **Original publication** | <sup>α</sup>0.83<sup>30</sup> | <sup>β</sup>0.96<sup>23</sup> | <sup>γ</sup>0.75<sup>20</sup> | <sup>δ</sup>0.91<sup>32</sup> |

The confusion matrices (Fig. 2A) for zero-shot blood cell classification on the Acevedo data set show that current LVLMs tend to misclassify cells into a few dominant classes: GPT-4o, Gemini, and LLaMA predominantly assigned cells to lymphocytes or segmented neutrophils, while LLaVA-Med consistently classified cells as bands or segmented neutrophils. The computational pathology model CONCH mostly classified cells as segmented neutrophils or myelocytes.

**Fig. 2.**
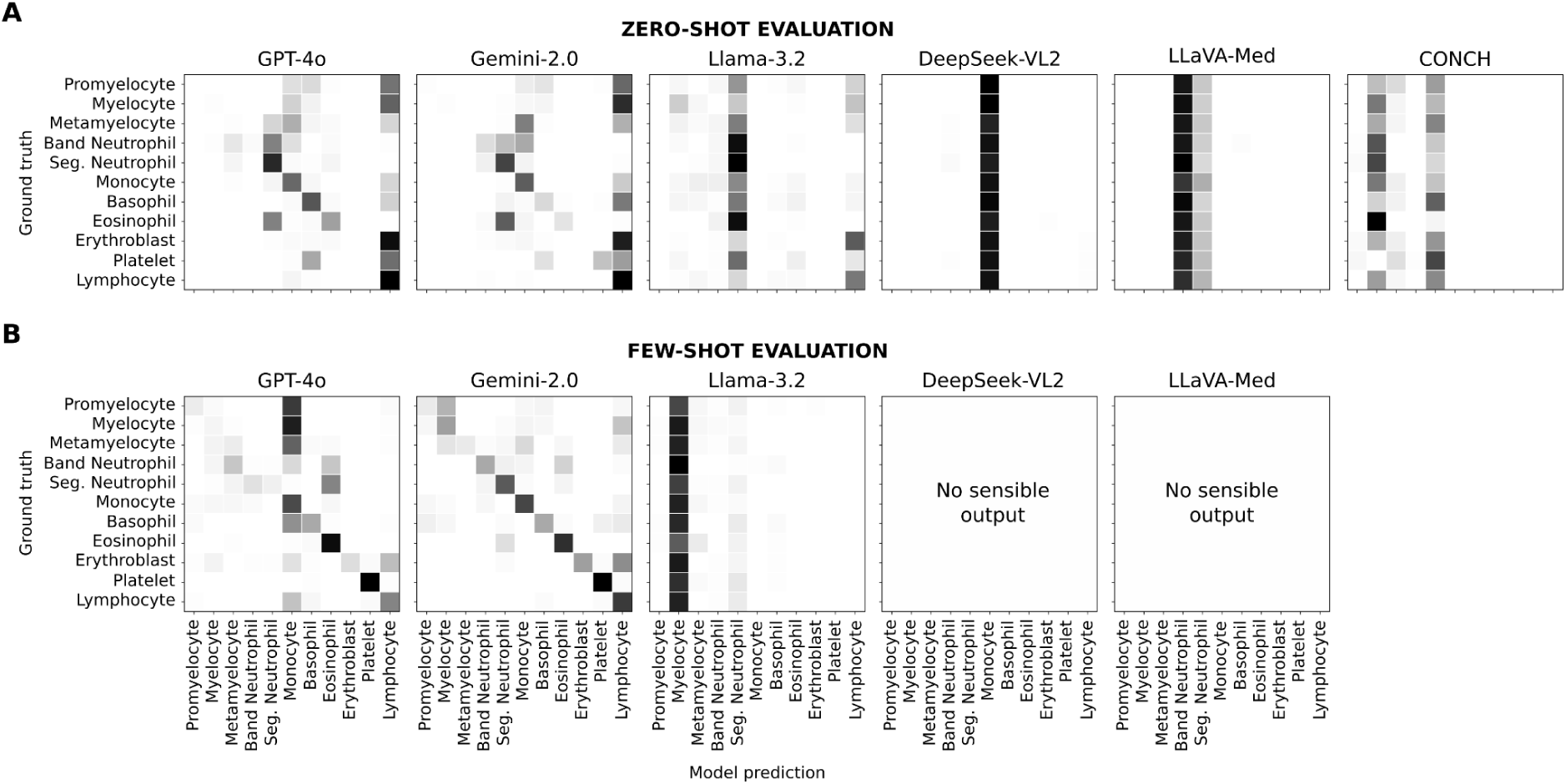
Large visual language models tend to classify cells into only a few cell types. Confusion matrices of models tested on the Acevedo dataset show high mis-classification in zero-shot evaluation (**A**) and moderate improvement for GTP-4o and Gemini-2.0 in few-shot (**B**) evaluation. Here, DeepSeek-VL2 generated erroneous text composed of random numbers and letters, while LLaVA-Med either gave no response or simply noted the presence of red blood cells and described their function.

Few-shot learning improved the performance, nearly doubling the scores for GPT-4o and Gemini across most tasks (Table 1). For instance, Gemini’s bone marrow cell classification score increased to 0.20±0.02, while its cervical smear classification score improved to 0.15±0.01. However, the scores remain closer to random model performance and still far from the state-of-the-art results. Fig. 2B visualizes the improved confusion matrices for blood cell classification in the Acevedo data. Few-shot learning led to more diagonally aligned confusion matrices for GPT-4o and Gemini-2.0, indicating better classification. LLaMA-3.2 performed worse in few-shot learning, yielding even lower accuracy than in the zero-shot setting. DeepSeek and LLaVA-Med did not produce sensible output in a few-shot setting.

We fine-tuned GPT-4o and DinoBloom on the Acevedo blood cell classification task, varying the number of training images per class, and evaluated performance on a left out test set (Fig. 3A). Model performance improved rapidly with increasing dataset size: With 10 images per class, GPT-4o’s weighted F1 score rose from 0.22±0.02 (no fine-tuning) to 0.55±0.06, further improving to 0.65±0.05 with 50 images per class, to 0.74±0.04 with 100 images per class, and plateauing form there on (Fig. 3A). Interestingly, a simple MLP trained on DINOv2 and DinoBloom models learns significantly faster and better than GPT-4o. In particular the hematology foundation model DinoBloom^29^ excels, achieving scores of 0.79, 0.86, and 0.87 for 10, 50, and 100 images per class, outperforming GPT-4o at every stage (Fig. 3A). The confusion matrices for fine-tuned GPT-4o and DinoBloom reveal that misclassifications primarily occur between morphologically similar cell types, such as myelocytes vs. metamyelocytes or segmented vs. band neutrophils—categories that are inherently difficult to distinguish (Fig. 3B).

**Fig. 3.**
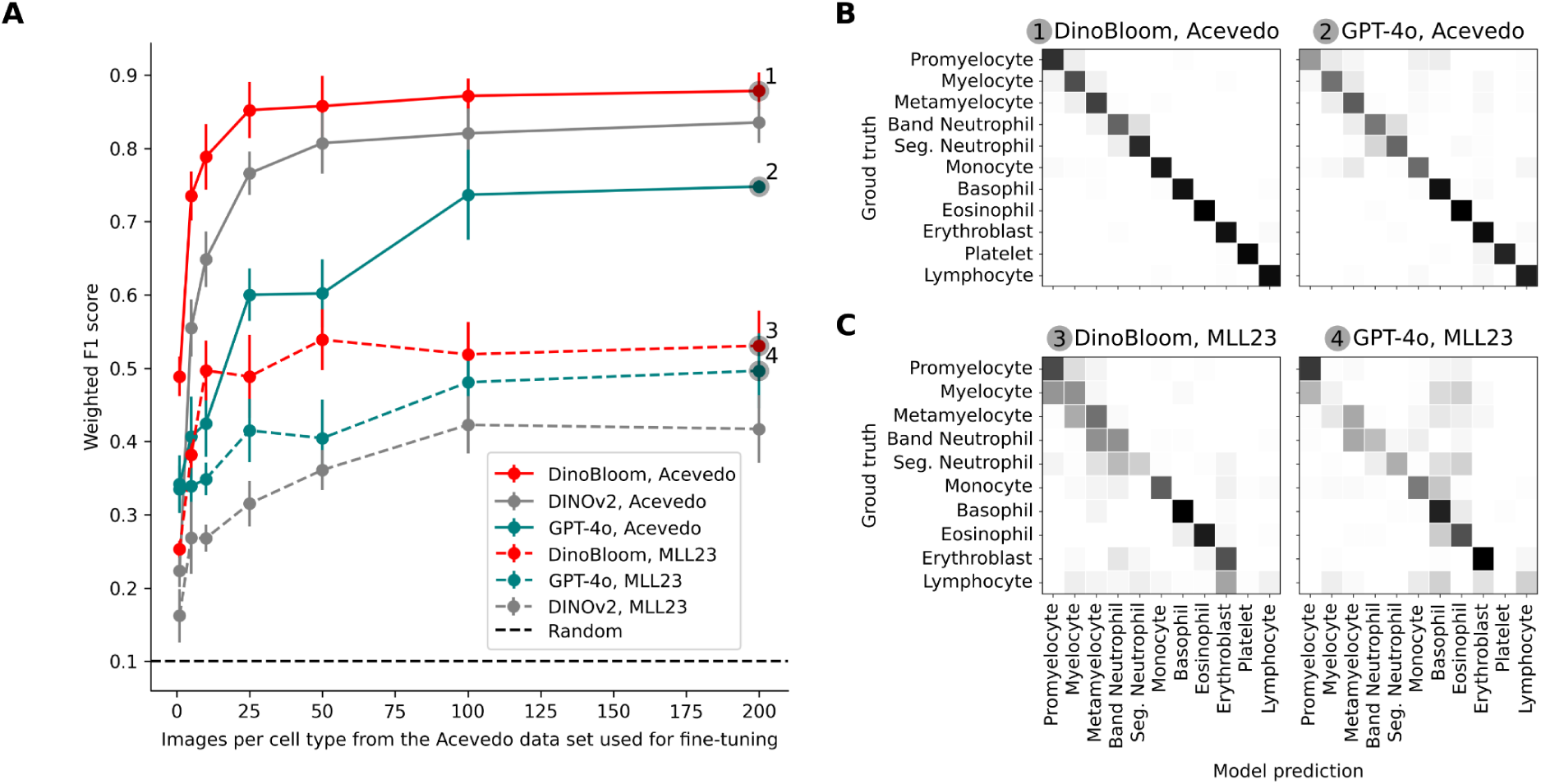
Domain-specific models outperform large vision language models. **A.** DinoBloom and DINOv2 models outperform GPT-4o at any fine-tuning dataset size in terms of weighted F1 scores in-domain (Models evaluated on an independent Acevedo test set, solid lines) and out-of-domain (MLL23 test set, dashed line). Confusion matrices of fined-tuned models evaluated on Acevedo (**B**) and MLL23 (**C**) prove superior performance of the domain-specific DinoBloom model.

To assess model generalizability, we evaluated the fine-tuned GPT-4o and DinoBloom on the out-of-distribution MLL23 test set (Fig. 3A, C). Notably, both models were fine-tuned for Acevedo cell classification. DinoBloom, with an MLP head, consistently outperformed GPT-4o across all training data sizes. At n=200, it achieved a weighted F1 score of 0.54, compared to 0.50 for the fine-tuned GPT-4o. The zero-shot GPT-4o classification performance on MLL23 was low at 0.19, only slightly above the random baseline of 0.10. Confusion matrices reveal that misclassifications mostly occurred between cell types that are morphologically close to each other (Fig. 3C).

What degree of explainability do LVLMs provide? Are they able to explain what morphological features they use to classify blood cells? We asked the models to classify all cells in the test set and score the importance of 19 morphological features (see Methods for details and the exact prompts used). We find cell type specific feature signatures (e.g. all models agreed on the importance of the ‘Nuclear Shape’ feature, Fig. 4A), and moderate differences between models. Some of the important features were indeed meaningful for the corresponding cell types, e.g. ‘Nuclear Segmentation’ for segmented neutrophils, ‘Cell Size’ for platelets, or ‘Chromatin Pattern’ and ‘Cell Size’ for eosinophils. However, important features used by morphological experts were also missed by the models, for example ‘Granule Presence’ and ‘Granule Type’ for eosinophils.

**Fig. 4.**
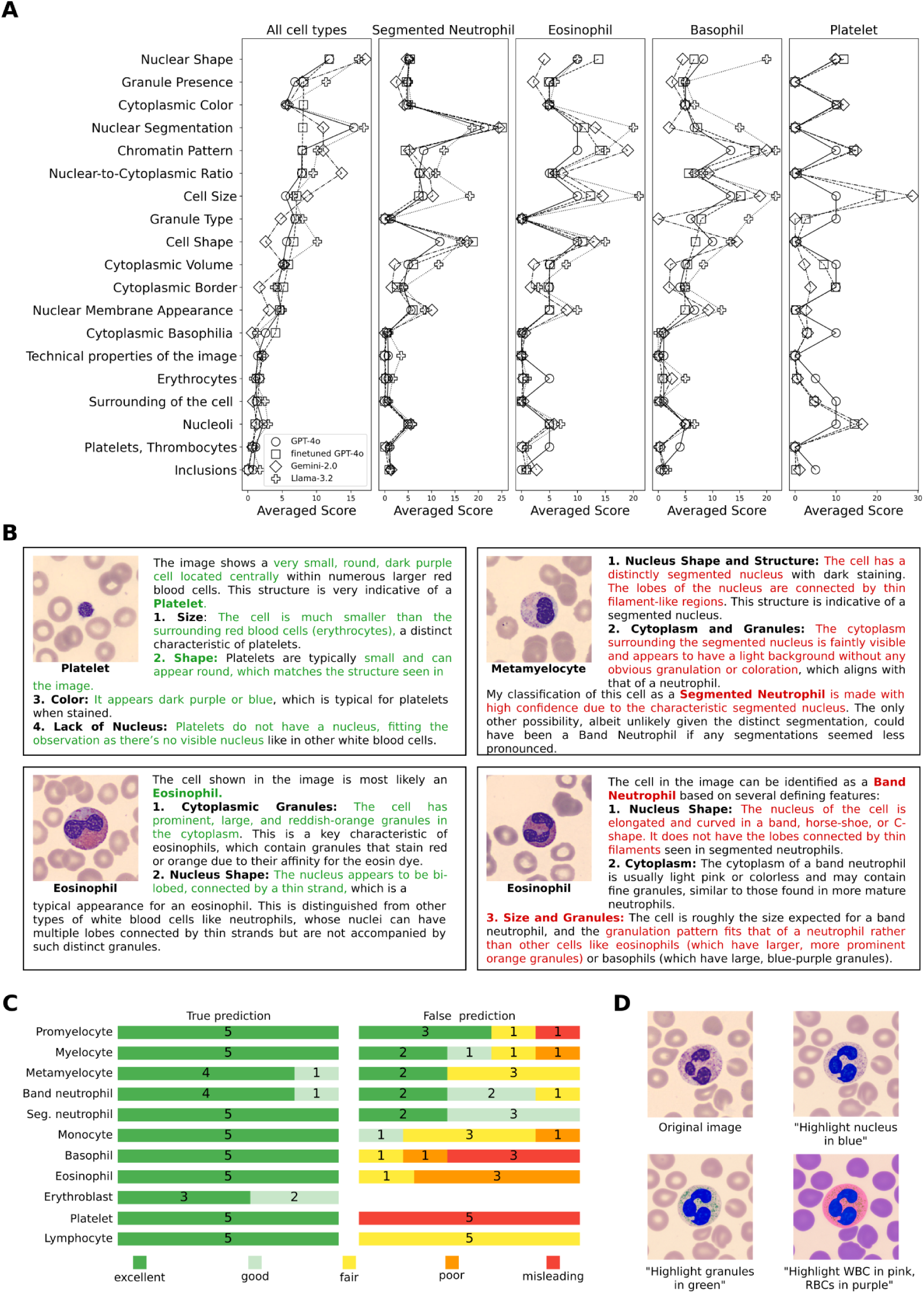
GPT-4o provides textbook explanations instead of truly interpreting cellular properties. **A.** Models assigned high importance scores to relevant morphological features. **B.** Free text decision explanations from the fine-tuned GPT-4o (two correct predictions right, tow wrong predictions left). **C.** An expert cytologist finds mostly excellent explanations for true predictions, but poor or misleading explanations in 30% of false predictions. **D.** Fine-tuned ChatGPT-4o segments cellular components successfully.

We next asked an expert cytomorphologist to assess free text explanations for 10 images per cell type (5 correctly and 5 incorrectly classified by the best performing LVLM, the fine-tuned GPT-4o, see Fig. 4B for examples) on the following scale: 1 - excellent, 2 - good, 3 - fair, 4 - poor, 5 - misleading. Explanations for correctly classified cell images received an average score of 1.1±0.3 (mean±s.d., n=55), while wrongly classified images received a considerably lower score of 2.8±1.4, with 30% of all explanations being rated as poor or misleading. Explanations of wrong classifications often missed or misinterpreted white blood cell features, e.g. recognizing uni-lobal nuclei as segmented, or describing cytoplasmic granulation incorrectly (Fig. 4B). The model often seemed to provide textbook descriptions of predicted cell types, regardless of the actual features present in the images. To evaluate GPT-4o’s vision capabilities, we asked the model to perform step-by-step segmentations (Fig. 4D). GPT-4o demonstrated a good understanding of cellular components by correctly identifying and segmenting relevant structures, but occasionally struggled with cell types such as platelets, basophils and erythroblasts (Supplementary Fig. 1). The model was unable, however, to highlight which parts of the cell were relevant for the cell classification.

In the free text answers, the models were also asked how certain they were about their classification decision. GPT-4o, fine-tuned GPT-4o and Gemini-2.0 were very confident about their decisions even when the predicted classes were wrong (Fine-tuned GPT-4o: “**Certainty Level**: High certainty that this is a segmented neutrophil because of the classical lobulated and segmented nuclear appearance.” Gemini-2.0: “I am quite certain (90-95%) that this is a Lymphocyte.”). Llama-3.2 was more realistic: “I am moderately certain about this classification. The image is not clear enough to confirm all of the features of a myelocyte, but the overall shape and size of the cell suggest that it is an immature granulocyte.” We conclude that the models are in general overestimating their reliability.

## Discussion

Our results demonstrate that current LVLMs substantially underperform in cytomorphology—a domain that, in many respects, poses simpler visual challenges than for example pathology. Across four clinically relevant cytomorphology tasks, generalist LVLMs often produced results close to random guessing, particularly in zero-shot settings. Models frequently defaulted to dominant classes such as segmented neutrophils and lymphocytes, which together represent over 75% of white blood cells in peripheral blood (Supplementary Table 1). This suggests reliance on prior probability rather than true morphological interpretation. Few-shot prompting improved performance, but it remained well below that of domain-specific models reported in the literature. The observed limitations likely reflect a lack of cytomorphology-specific data in the training corpora of LVLMs.

Fine-tuning GPT-4o yielded notable gains, but performance remained consistently inferior to a simple multilayer perceptron (MLP) trained atop a cytomorphology-specific foundation model. Importantly, domain-specific models not only achieved higher accuracy but also converged faster and required less computational effort. These models reached performance plateaus with as few as 100 images per class, aligning with prior findings^41,42^. Our findings also fit to the result of Jiang et al.^6^, who reported below-baseline performance for endoscopy images, chest x-ray, and skin lesions.

We also investigated explainability—a critical requirement for clinical deployment. Although LVLMs could generate plausible justifications, their explanations for misclassified cases revealed a significant limitation: they tended to provide generic, textbook-level descriptions rather than specific morphological reasoning grounded in the actual image features. This suggests weak integration between visual and language modalities, likely stemming from insufficient multimodal training on cytomorphology data. Future research should explore whether expanded training datasets or enhanced reasoning architectures can address these limitations and achieve the level of precise, image-specific explanations necessary for clinical practice.

In summary, current LVLMs remain unsuitable for high-stakes clinical applications in cytomorphological diagnostics. Their limitations in performance, generalization, and interpretability significantly lag behind purpose-built, domain-specific models. For clinical institutions and AI companies alike, investing in specialized models trained on carefully curated datasets offers a more effective and cost-efficient path forward. Our findings underscore three critical requirements for clinical readiness: improved multimodal alignment to better integrate visual and textual understanding, strategic inclusion of domain-specific data during pretraining, and rigorous evaluation protocols that reflect real-world diagnostic challenges. Until these fundamental issues are addressed, LVLMs cannot be considered viable tools for clinical diagnostic use.

## Acknowledgements

We thank Emre Akbas (Middle East Technical University, Turkey) for valuable feedback on the manuscript.

## Competing interests

We declare no competing interests.

## Ethics approval

This study did not require ethical approval.

## Data availability

The data used in this study is publicly available.

HiCervix: https://zenodo.org/records/11081816

WBCAtt: https://github.com/apple2373/wbcatt/tree/main/submission

Acevedo: https://data.mendeley.com/datasets/snkd93bnjr/1

BMC: https://www.cancerimagingarchive.net/collection/bone-marrow-cytomorphology_mll_helmholtz_fraunhofer/

MLL23: https://zenodo.org/records/14277609

## Code Availability

The code used in this study is available at: https://github.com/marrlab/LVLM_cytomorphology_eval

## Authors’ contributions

IK and MFD performed the LVLM evaluation and analysis with CM. JS developed model explainability analysis methodology with IK, MFD and CM. MB and KSG provided expert guidance. CM supervised the project. All the authors contributed to the manuscript writing.

## Use of generative AI

We evaluated generative AI models as described in the text in detail. We used OpenAI’s GPT-4o model for optimizing code and English text.

## Supplementary Material

### Prompts

#### Zero-shot and few-shot evaluation

Except for the CONCH model, we used prompts to get answers from LVLMs. For each dataset, we used following question prompts in zero-shot and few-shots settings:

***HiCervix:*** *“Consider the input image. Take a moment to think. Consider what features do cells in the image have. Which of the types listed below is shown?*

*Write just the cell type and nothing else. Choose one of the possible labels provided below (exactly as written here):*

*Normal cell*
*Endocervical cell*
*Repair cell*
*Metaplasia cell*
*Pseudokoilocytes by glycogen*
*Atrophy*
*Endometrial cell*
*Hyperchromatic crowded groups*
*Atypical squamous cells of undetermined significance*
*Low-grade squamous intraepithelial lesion*
*Atypical squamous cells, cannot exclude high-grade squamous intraepithelial lesion*
*High-grade squamous intraepithelial lesion*
*Squamous cell carcinoma*
*Atypical glandular cell*
*Atypical glandular cell- not otherwise specified*
*Atypical glandular cell- favor neoplastic*
*Adenocarcinoma*
*Atypical glandular cell (endocervical cell)- not otherwise specified*
*Atypical glandular cell (endometrial cell)- not otherwise specified*
*Adenocarcinoma of endocervical cell*
*Adenocarcinoma of endometrial cell*
*Fungal organisms morphologically consistent with Candida spp.*
*Bacteria morphologically consistent with Actinomyces spp.*
*Trichomonas vaginalis*
*Cellular changes consistent with herpes simplex virus*
*Coccobacilli/Shift in flora suggestive of bacterial vaginosis”*

***Acevedo:*** *“Consider the input image. Take a moment to think. Consider what features do the cells in the image have. Which of the white blood cell types listed below is shown?*

*Write just the cell type and nothing else. Choose one of the possible labels provided below (exactly as written here):*

*Band Neutrophil*
*Basophil*
*Eosinophil*
*Erythroblast*
*Lymphocyte*
*Metamyelocyte*
*Monocyte*
*Myelocyte*
*Platelet*
*Promyelocyte*
*Segmented Neutrophil”*

***BMC:*** *“Consider the input image. Take a moment to think. Consider what features do the cells in the image have. Which of the white blood cell types listed below is shown?*

*Write your full considerations but conclude your reply with ‘Answer:’ and then write one of the possible labels provided below (exactly as written here):*

*Abnormal eosinophil*
*Artefact*
*Basophil*
*Blast*
*Erythroblast*
*Eosinophil*
*Faggott cell*
*Hairy cell*
*Smudge cell*
*Immature lymphocyte*
*Lymphocyte*
*Metamyelocyte*
*Monocyte*
*Myelocyte*
*Band neutrophil*
*Segmented neutrophil*
*Other cell*
*Proerythroblast*
*Plasma cell*
*Promyelocyte”*

***WBCAtt:*** *label: “Consider the input image. Take a moment to think. Consider what features do the cells in the image have. Which of the white blood cell types listed below is shown? Write just the cell type and nothing else. Choose one of the possible labels provided below (exactly as written here):*

*Neutrophil*
*Eosinophil*
*Basophil*
*Lymphocyte*
*Monocyte”,*

*cell_size: “Consider the white blood cell shown in the input image. Take a moment to think. What is the size of the white blood cell? Choose one of the possible labels provided below (exactly as written here):*
*big*
*small”,*

*cell_shape: “Consider the white blood cell shown in the input image. Take a moment to think. What is the shape of the white blood cell? Choose one of the possible labels provided below (exactly as written here):*
*round*
*irregular”,*

*nucleus_shape: “Consider the white blood cell shown in the input image. Take a moment to think. What is the shape of the nucleus of the white blood cell? Choose one of the possible labels provided below (exactly as written here):*
*unsegmented-band*
*unsegmented-round*
*segmented-*
*segmented-bilobed*
*irregular*
*unsegmented-indented”,*

*nuclear_cytoplasmic_ratio: “Consider the white blood cell shown in the input image. Take a moment to think. What is the ratio of the nucleus to the cytoplasm of the white blood cell? Choose one of the possible labels provided below (exactly as written here):*
*low*
*high”,*

*chromatin_density: “Consider the white blood cell shown in the input image. Take a moment to think. What is the chromatin density of the white blood cell? Choose one of the possible labels provided below (exactly as written here):*
*densely*
*loosely”,*

*cytoplasm_vacuole: “Consider the white blood cell shown in the input image. Take a moment to think. Does the white blood cell have a cytoplasmic vacuole? Choose one of the possible labels provided below (exactly as written here):*
*no*
*yes”,*

*cytoplasm_texture: “Consider the white blood cell shown in the input image. Take a moment to think. What is the texture of the cytoplasm of the white blood cell? Choose one of the possible labels provided below (exactly as written here):*
*clear*
*frosted”,*

*cytoplasm_colour: “Consider the white blood cell shown in the input image. Take a moment to think. What is the colour of the cytoplasm of the white blood cell? Choose one of the possible labels provided below (exactly as written here):*
*light blue*
*blue*
*purple blue”,*

*granule_type: “Consider the white blood cell shown in the input image. Take a moment to think. What is the type of the granules in the white blood cell? Choose one of the possible labels provided below (exactly as written here):*
*small*
*round*
*coarse*
*nil”,*

*granule_colour: “Consider the white blood cell shown in the input image. Take a moment to think. What is the colour of the granules in the white blood cell? Choose one of the possible labels provided below (exactly as written here):*
*pink*
*purple*
*red*
*nil”,*

*granularity: “Consider the white blood cell shown in the input image. Take a moment to think. Does the white blood cell have granules? Choose one of the possible labels provided below (exactly as written here):*
*yes*
*no “*

For few-shot experiments, we presented an example from each class before asking the same question as in the zero-shot prompt.

#### Reviewing answers

Even though the models were asked to answer with the predicted class only, often they replied with a long answer justifying their decision. For example, Llama-3.2 answered:

*Considerations:*

1. *\*\*Nucleus Appearance**: The image shows cells with dark-stained nuclei. The shape and segmentation of nuclei are important for identifying white blood cell types*.
2. *\*\*Cytoplasm and Granules**: Observe the characteristics of the cytoplasm and whether it contains granules or not. Basophils and eosinophils typically have granules, but their color and dispersion differ*.
3. *\*\*Cell Size and Shape**: Consider the overall size and shape of the cells. Monocytes are generally larger with indented nuclei, while lymphocytes are smaller with a round nucleus*.
4. *\*\*Granule Color**: The color and size of granules can help distinguish between eosinophils, which have large red-orange granules, and basophils, which have bluish-purple granules*.
5. *\*\*Nuclear Segmentation**: Neutrophils have segmented nuclei, while blasts have larger, less segmented nuclei*.

*Based on these considerations, the cells in the image display characteristics consistent with basophils, such as a lobed nucleus obscured by dark granules*.

To extract the model’s predicted class, we used GPT-4o with the following prompt:

“*Chatbot answered: {chatbot’s answer}*

*Which of the classes listed below does the chatbot’s answer regarding the cell type belong to? Write just the label (exactly as written below) and nothing else:*

*NA (Chatbot is unsure/ambiguous/doesn’t know/no answer provided/class cannot be determined)*
*Band Neutrophil*
*Basophil*
*Eosinophil*
*Erythroblast*
*Lymphocyte*
*Metamyelocyte*
*Monocyte*
*Myelocyte*
*Platelet*
*Promyelocyte*
*Segmented Neutrophil”*

#### Quantitative feature importance

To quantify the importance of morphological features for the model’s decision-making, we used the following prompt:

*“Consider the input image. Take a moment to think. Consider what features do the cells in the image have. Which of the white blood cell types listed below is shown? Write just the cell type and nothing else. Choose one of the possible labels provided below (exactly as written here): Band Neutrophil, Basophil, Eosinophil, Erythroblast, Lymphocyte, Metamyelocyte, Monocyte, Myelocyte, Platelet, Promyelocyte, Segmented Neutrophil*.

*Now consider the cell features listed below. Think how much each of them contributed to your cell classification decision that you made above. Next to each feature, write an importance score, how much the feature was important for your classification decision. The scores should be float numbers. All the scores together should sum to 100*.

*Cell Shape, Cell Size, Nuclear Shape, Nuclear Segmentation, Nuclear-to-Cytoplasmic Ratio, Nuclear Membrane Appearance, Nucleoli, Chromatin Pattern, Cytoplasmic Volume, Cytoplasmic Color, Cytoplasmic Border, Granule Presence, Granule Type, Inclusions (Presence of Auer rods, Döhle bodies, or other cytoplasmic inclusions), Cytoplasmic Basophilia, Erythrocytes, Platelets, Thrombocytes, Surrounding of the cell, Technical properties of the image (resolution, light, noise, etc.)*.

*Are there any other features that you consider important for the classification decision? If yes, write them below.”*

#### Reasoning

We assessed the model’s reasoning ability using the following prompt:

*“Consider the input image. Take a moment to think. Consider what features do the cells in the image have. Which of the white blood cell types listed below is shown? Choose one of the possible labels provided below (exactly as written here): Band Neutrophil, Basophil, Eosinophil, Erythroblast, Lymphocyte, Metamyelocyte, Monocyte, Myelocyte, Platelet, Promyelocyte, Segmented Neutrophil*.

*Explain in detail your decision and the reasoning that lead you to the decision. Which parts of the cells and features did you consider? How certain are you about your classification? Which other labels could be correct? Why did you choose this label in the end?”*

#### Computer vision

To test the model’s segmentation abilities, we used the following prompts in sequence:

1. If present, highlight nucleus by painting in blue
2. Now, if present, highlight granules by painting in green
3. Now, highlight whole white blood cell in pink and all red blood cells in purple

### Fine-tuning

OpenAI allows the user to choose the following hyperparameters when fine-tuning GPT-4: batch size, learning rate multiplier, number of epochs. The user can also choose that the model selects the hyperparameters automatically. Experimenting with the settings, we found that the automatic option usually gives the best result. Batch sizes were typically 1-4, LR multipliers 1-2, and number of epochs 3-5. The entries in the .jsonl files that we used for fine-tuning had the following form:

*{“messages”: [{“role”: “system”, “content”: “You are an assistant that identifies cell types.”}, {“role”: “user”, “content”: “Consider the input image. Take a moment to think. Consider what features do the cells in the image have. Which of the white blood cell types listed below is shown? \n Write just the cell type and nothing else. Choose one of the possible labels provided below (exactly as written here):\n Band Neutrophil\n Basophil\n Eosinophil\n Erythroblast\n Lymphocyte\n Metamyelocyte\n Monocyte\n Myelocyte\n Platelet\n Promyelocyte\n Segmented Neutrophil”}, {“role”: “user”, “content”: [{“type”: “image_url”, “image_url”: {“url”: “https://[…]/image_43.jpg”}}]}, {“role”: “assistant”, “content”: “Band Neutrophil”}]}*

For fine-tuning DinoBloom-S, we added to the model a two-layer multilayer perceptron with a ReLU activation between layers and a hidden layer of 128 units. During the fine-tuning, the MLP was trained while the DinoBloom-S was kept frozen. We used the Adam optimizer with a learning rate of 1e-3. The model was trained for 100 epochs. We selected the model with the best validation loss. Training and evaluation was conducted on a Tesla V100 32GB GPU.

#### Random model

To calculate random performance, we generated 1,000,000 random ground truth labels, maintaining the original class distribution as weights for each dataset. Each random performance experiment was repeated 1,000 times.

### User experience

The first important information that we got when running the models was the user experience.

The best of all was GPT. The website interface is transparent and easy to use. After obtaining an API key and topping up the account, the model ran smoothly and fast. A downside of GPT is that the online instructions are scarce and chatGPT has not been trained on the most recent information on GPT API calling, so it cannot provide instructions.

Second best model in terms of user experience was Llama. Once it was set up, it ran smoothly. The downside is that it did not run on GPUs with less than 80GB of memory, in our case we needed an A100 GPU. It was still the slowest model of all of them. Few-shot evaluation was particularly slow, and it took around 12h for a dataset to complete. Another flaw that we noticed with Llama was that it always gave a very long answer with all the reasoning explained, even if it was asked to provide a short answer with the cell class only.

The third in user experience was Gemini. The web interface is in our opinion confusing and hard to navigate. The model would sometimes interrupt the runs with “resources exhausted” message. This is because Gemini limits the number of API calls per minute, with no clear information available about what the limits are. Increasing the sleep time between consecutive API requests to 5 s helped, but the runs would sometimes still get interrupted. So we ended up wasting a lot of time to rerun the Gemini jobs.

LLaVA-Med and DeepSeek VL2 required running on older versions of Cuda and PyTorch and older versions of Conda environments that are not easy to reconstruct. This took a significant amount of time to set up. DeepSeek-VL2 and LLaVA-Med failed to produce sensible outputs in the few-shot setting—DeepSeek-VL2 generated erroneous text composed of random numbers and letters, while LLaVA-Med either gave no response or simply noted the presence of red blood cells and described their function. CONCH worked smoothly but produced underwhelming results. DinoBloom worked flawlessly, but we used it only for the fine-tuning benchmark.

Fine-tuning with GPT requires preparing the data set in a form of a .jsonl file first and uploading the images to some web location. We found the instructions how to prepare the .jsonl files on the OpenAI website ^43^ too scarce, and it took a lot of trial and error to figure out how to prepare them correctly. Fine-tuning works in two steps. The user first uploads the data sets and after sets up the training job. The .jsonl files do get checked during the uploading but even if they get approved, there is often an error with the .jsonl file reported later while running the training. Some of our single cell images were rejected with an explanation that they “contain faces” or “contain captchas” which “violates Open AI” policies. The model creates and stores a few checkpoints during the training. It was slightly surprising that the checkpoints were not created in the steps when the validation loss was the lowest, but they seemed to be created a bit randomly. It was also not clear whether the final result model was taken to be the one with the lowest loss. Slightly surprising for a rather expensive service - fine-tuning on a dataset consisting of 1100 images for 10 epochs costs around 100 €.

**Supplementary Figure 1.**
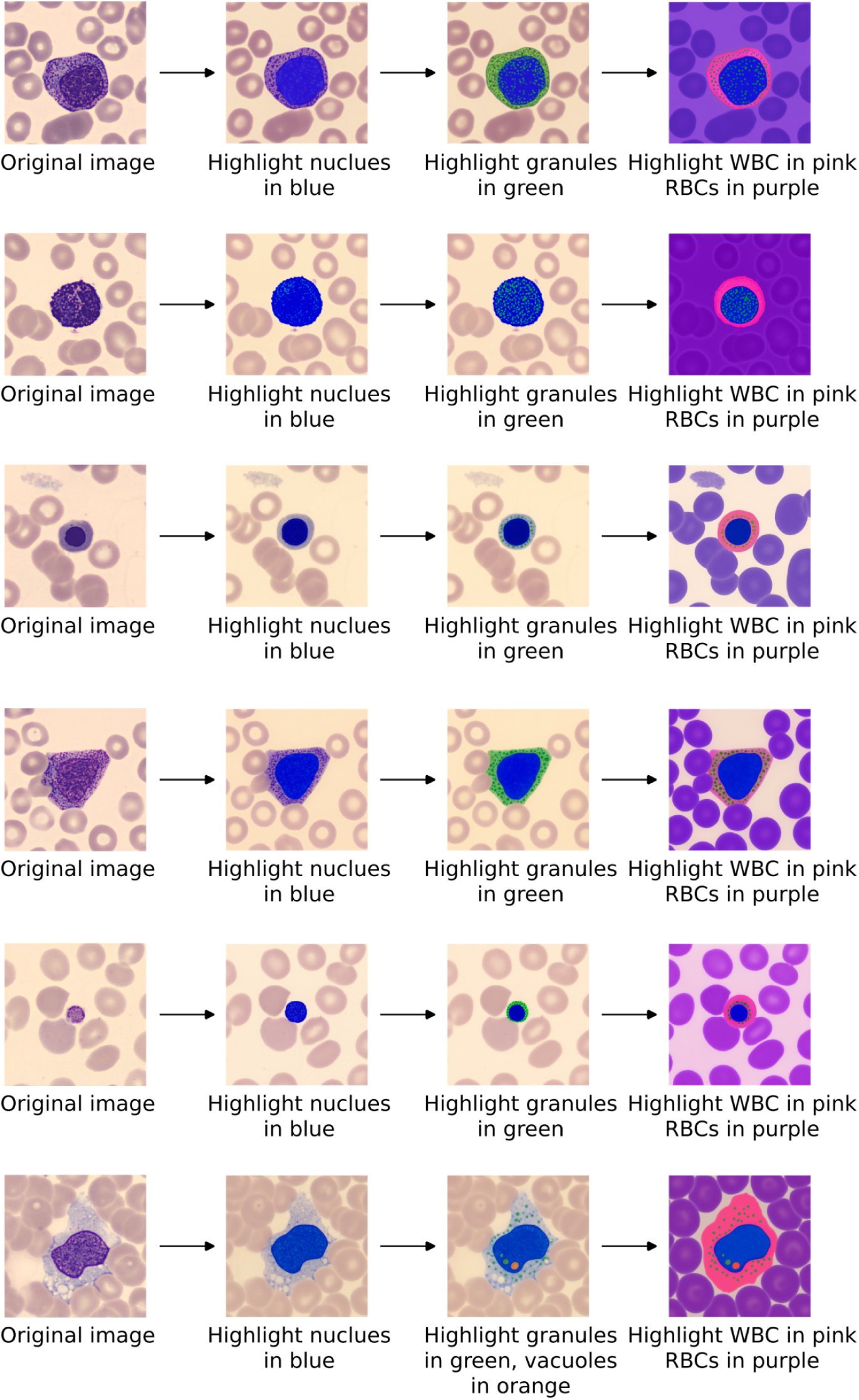
Step-wise cell segmentation using GPT-4o. Although the model was prompted to segment nucleus and granules only if present, it consistently segmented certain components regardless of their presence, e.g. segmenting granules in monocytes, where they are typically absent. The nucleus was mostly segmented correctly, except in cases where it is not actually present, such as in platelets (third row). As expected, the model fails to segment the nucleus in basophils, where it is usually obscured by coarse granules. Otherwise, segmentations appear accurate when clear nuclei, granules, and red blood cell structures are present.

**Supplementary Table 1.**
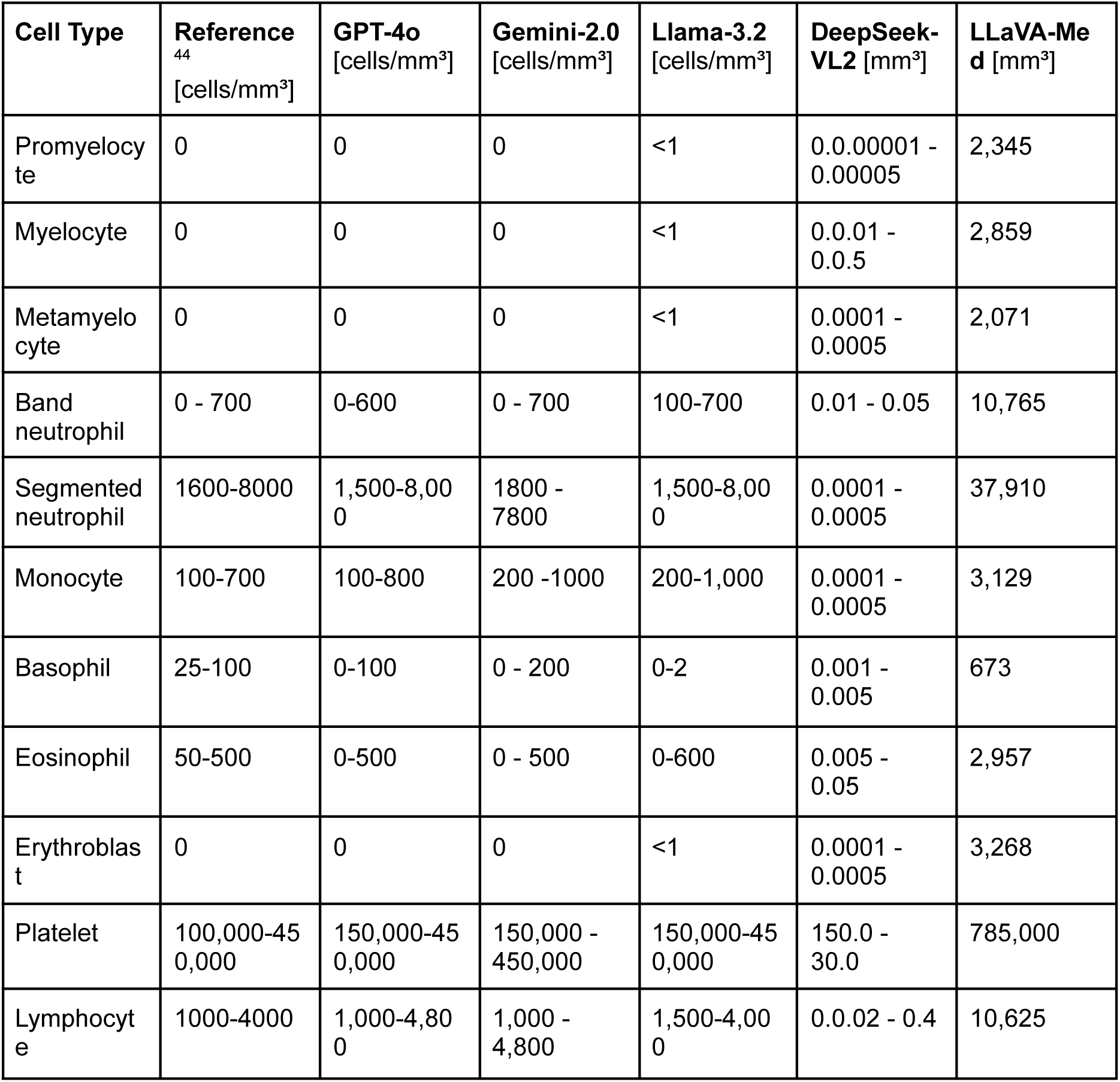
Blood cell type counts in peripheral blood smears as predicted by LVLMs. Responses are presented in their raw output format, including the units provided by the models.

## Notes

### Competing Interest Statement

The authors have declared no competing interest.

### Funding Statement

This study did not receive any funding

### Author Declarations

All data used in this study is publicly available: HiCervix comprises 40,229 cervical cells from 4,496 whole slide images, categorized into 29 classes. The HiCervix dataset includes normal epithelial cells, infectious agents, and malignant cells. https://zenodo.org/records/11081816 WBCAtt: contains morphology annotations for 10,300 images from the Acevedo data set31. Labels are provided for 11 fine-grained morphological attributes like nucleus shape, chromatin density, granularity, or cytoplasm texture. https://github.com/apple2373/wbcatt/tree/main/submission Acevedo: provide 17,092 white blood cell images from peripheral blood smears, labeled with 11 different cell type annotations. https://data.mendeley.com/datasets/snkd93bnjr/1 BMC: is a collection of 171,373 white blood cell images from bone marrow smears collected from 945 patients. The cells were expert-labeled into 21 different cell types. https://www.cancerimagingarchive.net/collection/bone-marrow-cytomorphology_mll_helmholtz_fraunhofer/ MLL23: was used only as an external test set for fine-tuned models. It includes over 40,000 expert annotated peripheral blood single cell images categorized into 18 classes. https://zenodo.org/records/14277609

